# The Impact of the COVID-19 Pandemic on Rabies Reemergence in Latin America: the case of Arequipa, Peru

**DOI:** 10.1101/2020.08.06.20169581

**Authors:** Brinkley Raynor, Elvis W. Díaz, Julianna Shinnick, Edith Zegarra, Ynes Monroy, Claudia Mena, Ricardo Castillo-Neyra

## Abstract

Over the past decades, there has been tremendous progress towards eliminating canine rabies in Latin America. Major components of rabies prevention programs in Latin America leading to these successes have been constant and intense surveillance for rabid dogs and uninterrupted yearly mass dog vaccination campaigns. However, vital measures to control COVID-19 in Latin America have had the negative trade-off of jeopardizing these rabies elimination and prevention activities. In this paper, we aimed to assess the effect of interrupting canine rabies surveillance and mass dog vaccination campaigns on rabies trends. We built a deterministic compartment model of dog rabies dynamics parameterized for conditions found in Arequipa, Peru, where there is an ongoing dog rabies epidemic. Our model suggests that a decrease in canine vaccination coverage as well as decreased surveillance leading to an increased length of survival of infected dogs could lead to a sharp rise in canine rabies and, subsequently, human rabies risk. We examined our results over the best estimate of the basic reproductive number in Arequipa (R_0_ = 1.44) and a range of plausible values for R_0_ (1.36 - 2). The rising trend was consistent. It is very possible that COVID-19 will continue to challenge our public health departments in the short- and medium-term. Innovative strategies to conduct dog vaccination and rabies surveillance during these trying times should be considered to safeguard the achievements made in Latin America towards the elimination of dog-mediated human rabies.

## Resumen *(Abstract in Spanish)*

En las últimas décadas ha habido un tremendo progreso hacia la eliminación de la rabia canina en América Latina. Los principales componentes de los programas de prevención de la rabia en América Latina que condujeron a estos éxitos han sido la vigilancia constante e intensa de los perros con rabia y las campañas anuales de vacunación masiva ininterrumpida. Sin embargo, las medidas esenciales para controlar el COVID-19 en América Latina han tenido el balance negativo de poner en peligro estas actividades de prevención y eliminación de rabia. En este artículo, nuestro objetivo fue evaluar el efecto que la interrupción de la vigilancia de la rabia canina y las campañas de vacunación masiva de perros tendría en las tendencias de la rabia. Modelamos la dinámica de la rabia canina mediante un modelo determinístico de comportamientos parametrizado para las condiciones que se encuentran en Arequipa, Perú, donde hay una epidemia de rabia canina en curso. Nuestro modelo sugiere que una disminución en la cobertura de vacunación canina, así como una disminución en la vigilancia (que llevaría a una mayor supervivencia de los perros infectados), podría llevar a un aumento súbito de rabia canina y, seguidamente, del riesgo de rabia humana. Examinamos nuestros resultados sobre la mejor estimación del número reproductivo básico en Arequipa (R_0_ = 1.44) y un rango de valores plausibles para R_0_ (1.36 - 2). La tendencia al alza fue consistente. Es muy posible que el COVID-19 continúe desafiando a nuestros departamentos de salud pública a corto y mediano plazo. Por lo tanto, deben considerarse estrategias innovadoras para llevar a cabo la vacunación de perros y la vigilancia de la rabia durante estos tiempos difíciles para salvaguardar los logros alcanzados en América Latina hacia la eliminación de la rabia humana mediada por perros.

Palabras Clave:

COVID-19, vacunación masiva, modelo matemático, Una Salud, rabia, SARS-CoV-2, zoonosis.

## Introduction

During the last decades, enormous progress has been achieved towards the elimination of canine rabies in the Americas (*1*–*3*). By 2019, health authorities in the Americas felt that Latin America was closer than ever to achieving the elimination of human deaths by rabies. The Pan American Health Organization (PAHO) announced on the eve of World Rabies Day – 28 September that only five human cases were reported in the region in the previous 12 months (*4*). The number of people dying from rabies reduced by more than 30 fold in 25 years (*5*). This reduction in human cases followed a drastic reduction of reported rabid dogs in Latin America from 15,686 in 1982 to 1,131 in 2003 (*5*). Both achievements were due mainly to a coordinated regional plan that involved multi-pronged strategies and continuous activities conducted by national governments and local communities (*3*). The successes of this regional endeavor have continued until recent years (*6*). The COVID-19 pandemic caused by the SARS-CoV-2 coronavirus has disrupted these strategies and activities in Latin America and could jeopardize the elimination prospects in the whole region.

Among the different strategies to prevent human rabies around the world, the most effective is mass dog vaccination (*7*–*9*). In most Latin American countries, the Ministries of Health organize annual or biannual mass canine rabies vaccination campaigns in areas affected and unaffected by canine rabies (*3*). During the few days these campaigns are held every year, a massive movement of people is required: many vaccination teams are deployed to salient locations all over cities to set up vaccination tents, many other vaccination teams visit houses door-to-door in remote or inaccessible areas, health promotion workers use megaphones to advertise the campaign on foot or from open trucks, and most importantly, all dog owners are expected to leave their houses and take their dogs to the vaccination points (*3*, *10*). Risk of SARS-CoV-2 transmission and sequential efforts to minimize that risk are huge barriers for implementation of mass dog vaccination campaigns.

Even though the most cost-efficient and high impact intervention to prevent, control, and eliminate canine rabies is mass dog vaccination (*8*, *9*), surveillance is also a vital component of rabies control programs (*11*–*13*). In Latin America, since 1983, rabies control programs have included intense surveillance (*3*). Regionally, in Latin America data are collected in a system for epidemiological surveillance of rabies called SIRVERA (*3*, *14*) that allows centralization of data, sharing information, evaluating progress, identifying areas with inappropriate surveillance, and detecting anomalies (e.g. outbreaks, expansion). Nationally and locally, surveillance consists mostly of submitting brain samples representing a specific proportion of the dog population (0.2% of the dog population per year is considered ideal in Latin America (*3*), a quota system that does not necessarily inform rabies control and elimination programs (*15*). The surveillance system also seeks to increase awareness and facilitate communication between communities and health inspectors so citizens report suspect dogs (e.g. animals neurological signs, excessively aggressive, salivating profusely, changing mood, or dying suddenly).

In response to the report of a suspect or confirmed rabid dog, a veterinarian or health inspector eliminates any dogs exhibiting rabies signs and submits samples for diagnostic testing of rabies (*16*, *17*). Any submissions testing positive are further responded to with focus control activities around the location the dog was originally found. A team of public health professionals is deployed to the field to conduct broad control and prevention measures such as dog vaccinations, administration of post-exposure prophylaxis, and removal oscf exposed (bitten) dogs (*16*–*19*). The most direct effect of such activities on rabies virus transmission is the prompt removal of rabid dogs, which effectively reduces the infectious period, the time a rabid dog is able to transmit the rabies virus (*20*), and the elimination of in-contact dogs which could shorten the transmission chains (*21*).

SARS-CoV-2 has infected more than 5.1 million people in Latin America as of August 5, 2020 and the pandemic has required an unprecedented, coordinated effort among national public health departments (*22*–*24*). Public health departments have necessarily shifted their focus and resources to implementing stay-at-home orders and ramping up emergency preparedness efforts. Moreover, veterinarians and other authorities have considered dog vaccination a high risk activity during the pandemic or a non-essential veterinary activity (e.g. non-urgent or non-emergency care) (*25*). Changes in resource allocation as well as professional activities and public mobility may have impacts on the control and surveillance of other diseases. In the case of rabies in Peru, and other Latin American countries, the yearly mass dog vaccination campaigns, the cornerstone of rabies prevention, have been postponed and likely will be downsized this year. Surveillance and focus control efforts have also been scaled back due to the COVID-19 pandemic. In this paper, we use a deterministic compartment model to explore the long-term effects of short-term changes to the rabies prevention protocols that have been developed and maintained over the past three decades. Specifically, we investigate how a reduction in canine vaccination coverage, decreased rabies surveillance, and decreased focus control efforts can affect canine rabies dynamics in Arequipa, Peru.

## Methods

### Model description

We created a deterministic compartment model of canine rabies transmission in Arequipa, Peru. The model distributes the canine population between 4 population states-vaccinated (in yearly vaccination campaigns), susceptible, exposed (via the bite of a rabid dog) and infectious (Figure 1). Table 1 describes the parameters included in the model.

**Figure 1:**
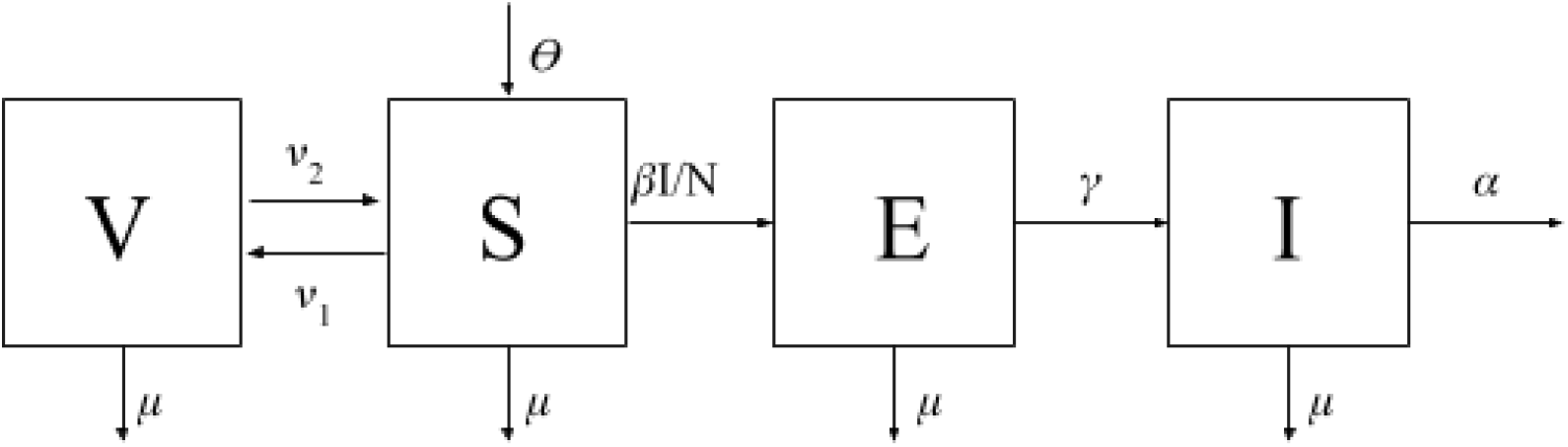
A flow diagram depicting 4 different population states (V-vaccinated, S-susceptible, E-exposed, I-infected) of dogs with arrows depicting the movement of individuals between states.

**Table 1:**
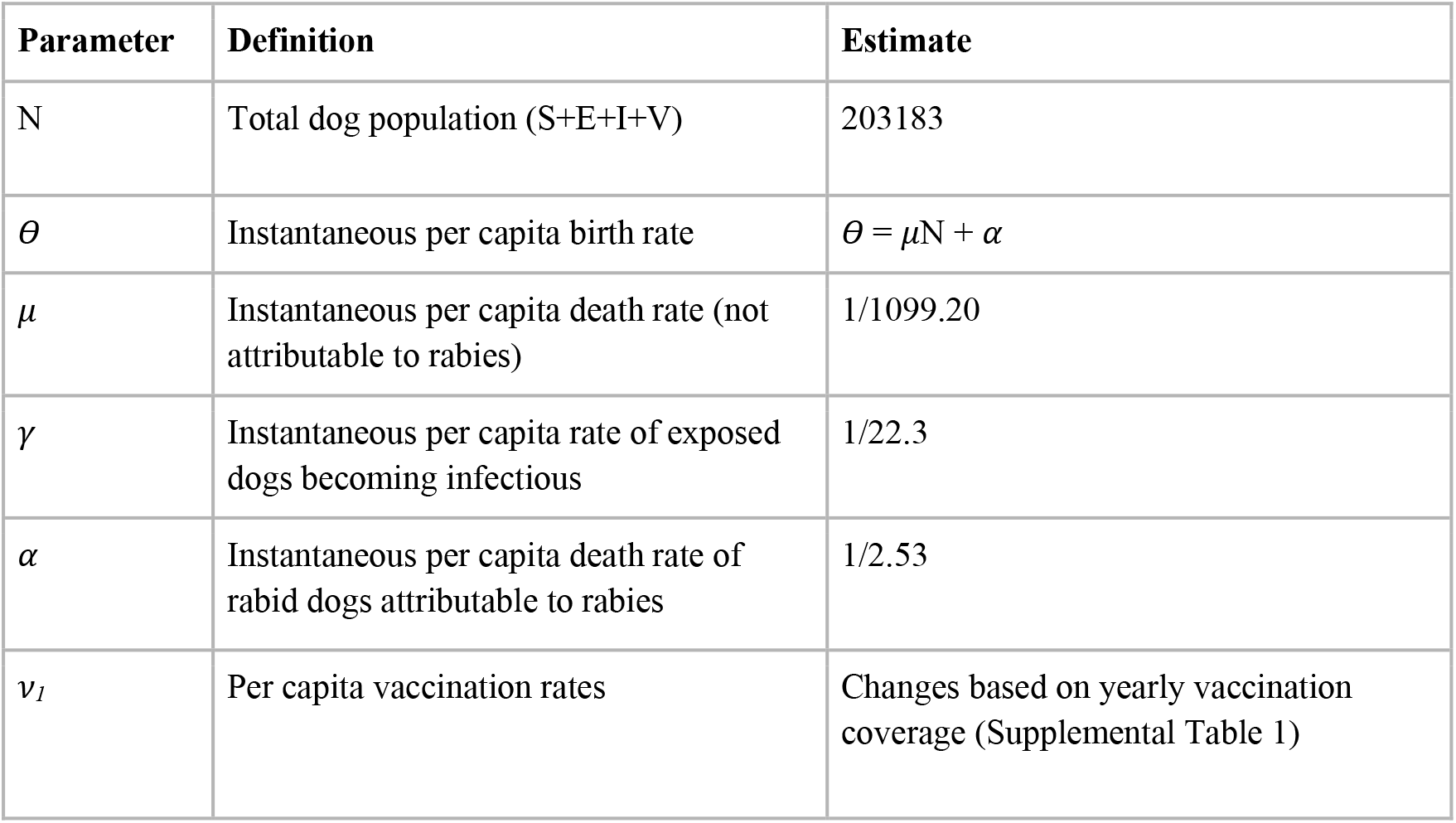
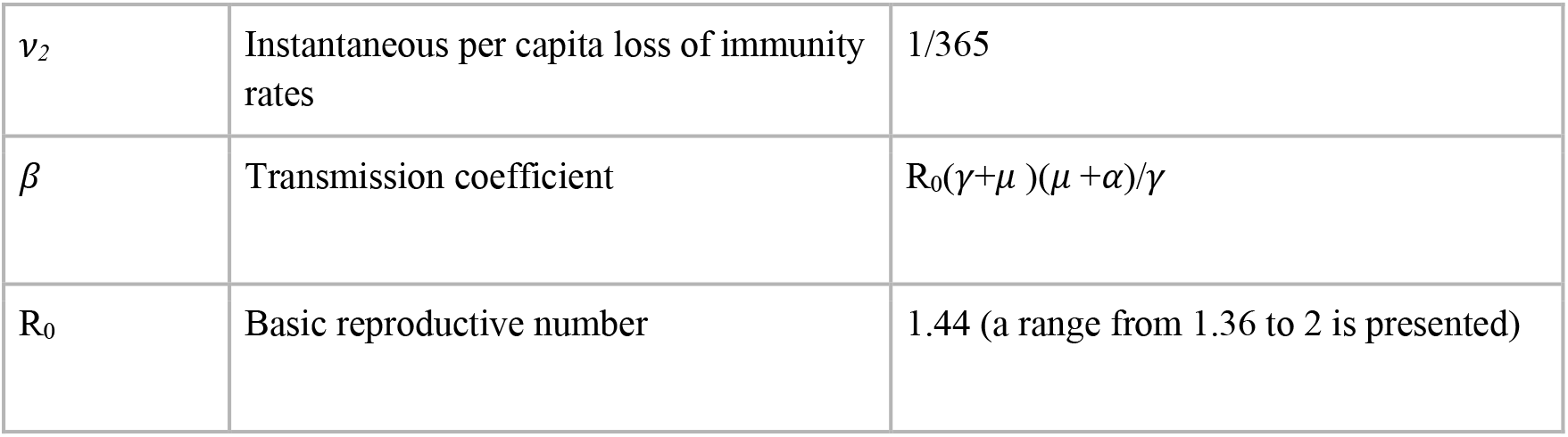
Definitions of parameters.

Equations depicting the movement between compartments can be expressed as:

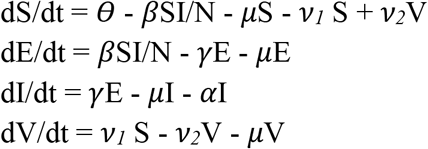

### Parameter estimation

The total population, N, was calculated based on estimates provided by the Peruvian Ministry of Health to be 203,183 dogs (*26*). The background death rate, *μ*, was calculated as the inverse of the average age of the dog population. Average age was calculated from a 2019 survey of over 3000 dogs to be 1099.2 days [1063.3, 1135.1]. Survey methods are described in detail elsewhere (*27*). The birth rate, *θ*, was calculated to maintain a steady state equilibrium: *θ* = *μ*N + *α*I.

The incubation period of rabies (1/*γ*) was assumed to be similar to rates reported in the literature. Hampson et al. found the maximum likelihood estimate of the mean incubation period from when exposed dogs were bitten to when they become infectious to be 22.3 days [20.0, 25.0] (*20*). The infectious period (1/*α*) when surveillance is in place, was estimated from Arequipa focus control data to be 2.5 days [1.9, 3.1] based on the difference between when owners reported their dog began showing symptoms and when they were euthanized by the public health veterinarians or health inspectors.

The rate of immunity loss (*v_2_*) is estimated to be the inverse of the immunity period offered by the vaccine. The vaccine used in the vaccination campaigns is rated for 1 year, so correspondingly *v_2_* = 1/365. Vaccination rates vary by year depending on coverage rates of the vaccination campaign. We make the simplifications that immunity is immediate upon vaccination and that the entire vaccination campaign happens in one day (though in reality it is spread across several days to weeks). In this way, immunity is “pulsed” once yearly and then immediately begins to decay. Supplement Table 1 shows estimated coverage rates from longitudinal survey data. Because vaccination coverage rates are based on a few days or a single day (the vaccination campaign), we assume a single day for our calculations (*10*).The pulsed vaccination rate (*v_1_*) can be calculated then as vaccination coverage = 1-e^-v1*t^, where t =1 day.

The transmission coefficient, *β*, can be very difficult to measure, but can be derived from the equation for R_0_. Using the next generation matrix methods, an equation for R_0_ can be derived from the disease system:

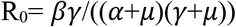

One challenge in estimating R_0_ for canine rabies globally is lack of accurate case counts. Rabies cases in humans are grossly underreported; in dogs, this trend is even more apparent (*3*, *6*, *28*–*32*). The WHO estimated that though in 2010 less than 10 human cases were reported in Latin America, there were in reality closer to 200 (*13*). In the literature the R_0_ for canine rabies across the world is reported from 1-2 (*20*, *33*–*35*).

In Arequipa, R_0_ was estimated from data collected from focus control team members responding to positive cases. Out of 214 cases, there are data on 33 cases about the number of secondary dogs that a rabid dog bit. Hampson et al. showed there is about a 0.49 probability that a dog will contract rabies if bitten by a rabid dog with a binomial confidence interval of [0.45, 0.52] (*20*). To estimate R_0_ for Arequipa, we randomly assigned a probability form the binomial distribution described by Hampson and used a bootstrap resampling method to estimate mean and 95% confidence intervals (*20*). We estimate R_0_ to be 1.36 [1.05, 1.88]. This R_0_ estimation should be considered as the low end of possible values as the data are biased due to being from focus control reports; these were cases that were responded to and controlled, limiting their number of secondary cases. Furthermore, we calculated R_0_ from *β* by fitting our simulated monthly incidence of infected dogs to the reported rabies case data using a least squares fitting approach (*36*, *37*). We use a conservatively low estimate of reporting rate (10%) of rabid dog cases. Due to the high number of people killing rabid dogs without reporting them and the high number of dogs hit by cars and never investigated, 10% is likely an overestimation of reporting rate for canine rabies cases in Arequipa. Using this rate of underreporting, we estimated an R_0_ of 1.44.

We present results for the possible range of R_0_ from 1.36, our low estimate from the focus control data, to 2, the high estimate from the literature, in order to show trends for a wide possibility of R_0_, the true value of which is unknown. However, we also present results specifically for R_0_=1.44, our best estimate for canine rabies in Arequipa.

### Interventions

Disruptions caused by COVID-19 will interrupt two key rabies elimination activities: mass dog vaccination and canine rabies surveillance. The disruption of each of these activities will affect several parameters in the model. Many vaccination programs around the world, not only against rabies, have been affected by scarce funds already shifted towards pandemic response and fear of being infected with the COVID-19 virus (*38*). Similarly, for canine rabies, the yearly vaccination campaigns are in jeopardy of being completely skipped this year in multiple countries, which would lead to an increased pool of susceptible dogs vulnerable to infection. To examine the effects of vaccination interruption in the model we change *v_1_*, the instantaneous per capita vaccination rate to reflect different scenarios: meeting the regional (*39*) and national goal of 80% coverage (*17*), a complete cancellation scenario of 0% coverage, and an intermediate effort of 58% coverage to match rates obtained previously (*27*).

Changes in city life during quarantine may also impact rabid dogs’ survival in several ways. First, fewer people are leaving home and, therefore, reports of rabid dogs have decreased to almost zero. Second, even if rabid dogs are reported, COVID-19 protocols have disrupted rabies response teams, so euthanization and removal of rabid dogs are delayed. Third, a decrease in traffic during the COVID-19 lockdown has led to increased survival time of disoriented dogs that otherwise would have been hit by cars (*40*). Hampson et al. found that rabid dogs died of the disease in an average of 3.7 days if they were not killed (*20*). Therefore, to examine the effect of increased survival time of rabid dogs due to COVID we shift the death rate (*α*) from a mean survival time of 2.5 days to 3.7 days.

### Computation

All computation was done using R (*41*).

## Results

We have reports of 214 rabid dogs in Arequipa since March 2015 (Figure S1). During the last four years 0.93 rabid dogs were reported per week, until March 2020. Since then, no rabid dogs have been reported due to the sharp drop in canine rabies surveillance. In recent months since COVID-19 control measures have been initiated, there has been a 10-fold decrease in samples submitted to be tested for rabies. However, the presence of a positive feline sample indicates that rabies virus is still circulating in the city. Our model has reasonable matching to the reported case data assuming a reporting rate of 10% and an R_0_ of 1.44 (Figure S1). The full dynamics (without any scaling for underreporting can be seen in Figure 2. Represented in Figure 2 is the cyclic nature of immunity and transmission caused by the yearly vaccination campaigns. Population immunity provided by the yearly vaccination campaign decays quickly due to high rates of population turnover (controlled by parameter *μ*) and loss of vaccine-provided immunity (controlled by parameter *ν_2_*) (Figure 2A). The proportion of the population affected by rabies transmission is so small that it is not apparent when shown together with the susceptible and vaccinated population (Figure 2A). However, the isolated exposed and infectious population dynamics follow a cyclic pattern (Figure 2B) caused by the pulses of immunity and subsequent decay: waves of exposed and infected dogs rise as population immunity falls. We also examined this behavior over a select range of possible values of R_0_ (Figure 2C) and though the amplitude of peaks may change, the rising trends remain the same. The trends are consistent for the full range of possible values of R_0_ from 1.36-2.0 (Figure S2).

**Figure 2:**
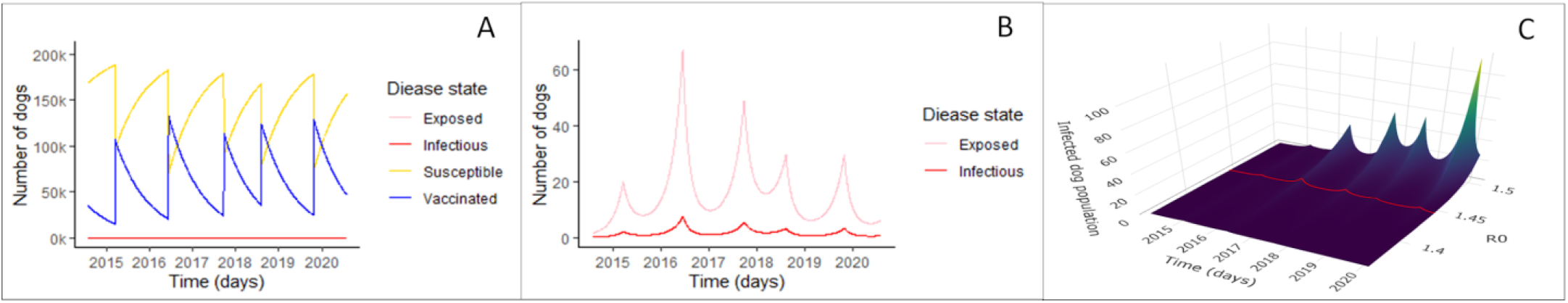
Rabies compartmental model results. Panel A shows the dynamics of all disease states in the best fit rabies model for Arequipa, Peru. The blue line shows the vaccinated dog population numbers over time and the yellow line shows the susceptible population dynamics. Because the proportion of rabies exposed (pink line) and infected dogs (red line) is so small, these dynamics are not apparent in Panel A. Panel B highlights these exposed and infectious dynamics with an adjusted scale. Panel C shows infected population dynamics for a range of R_0_ from 1.36-1.5. In other words, it represents the red line of infected population dynamics shown in B but for a range of R_0_. The trends extend for the full range of possible values of R_0_ [1.36, 2.0] which can be seen in supplementary materials (Fig S2).

Interventions are displayed both as a surface plot with the full range of possible values of R_0_, and as a 2D line plot with R_0_ = 1.44, representing our best estimate for Arequipa, Peru (Figure 3). We investigated the effects of changes in rabies prevention strategies including decreased vaccination campaign coverage represented by the parameter *v_1_* (Figure 3A-C, E) and decreased surveillance represented by the parameter *α* (Figures 4D,E). When vaccine coverage reaches the 80% recommended by PAHO, the numbers of infected dogs are suppressed to nearly 0 (Figure 3A). Conversely, with no vaccination coverage at all (due to a cancelled vaccination campaign, cases begin to grow exponentially (Figure 3C). However, even intermediate coverage (Figure 3B), or in other words, not hitting the 80% recommended by PAHO (Figure 3A) has a significant impact on suppressing the rise in infected numbers compared to no vaccination coverage at all (Figure 3C).

The effect of decreased surveillance and subsequent focus control is postulated to result in increased rabid dog survival time from 2.5 to 3.7 days as seen in Figure 3D - though incidence increases, the number of infected dogs can still be dampened by mass vaccination. The worst case scenario, where all control activities, mass dog vaccination, surveillance, and focus control, cease, results in a marked exponential rise in rabies cases within a few months (Figure 3E).

**Figure 3:**
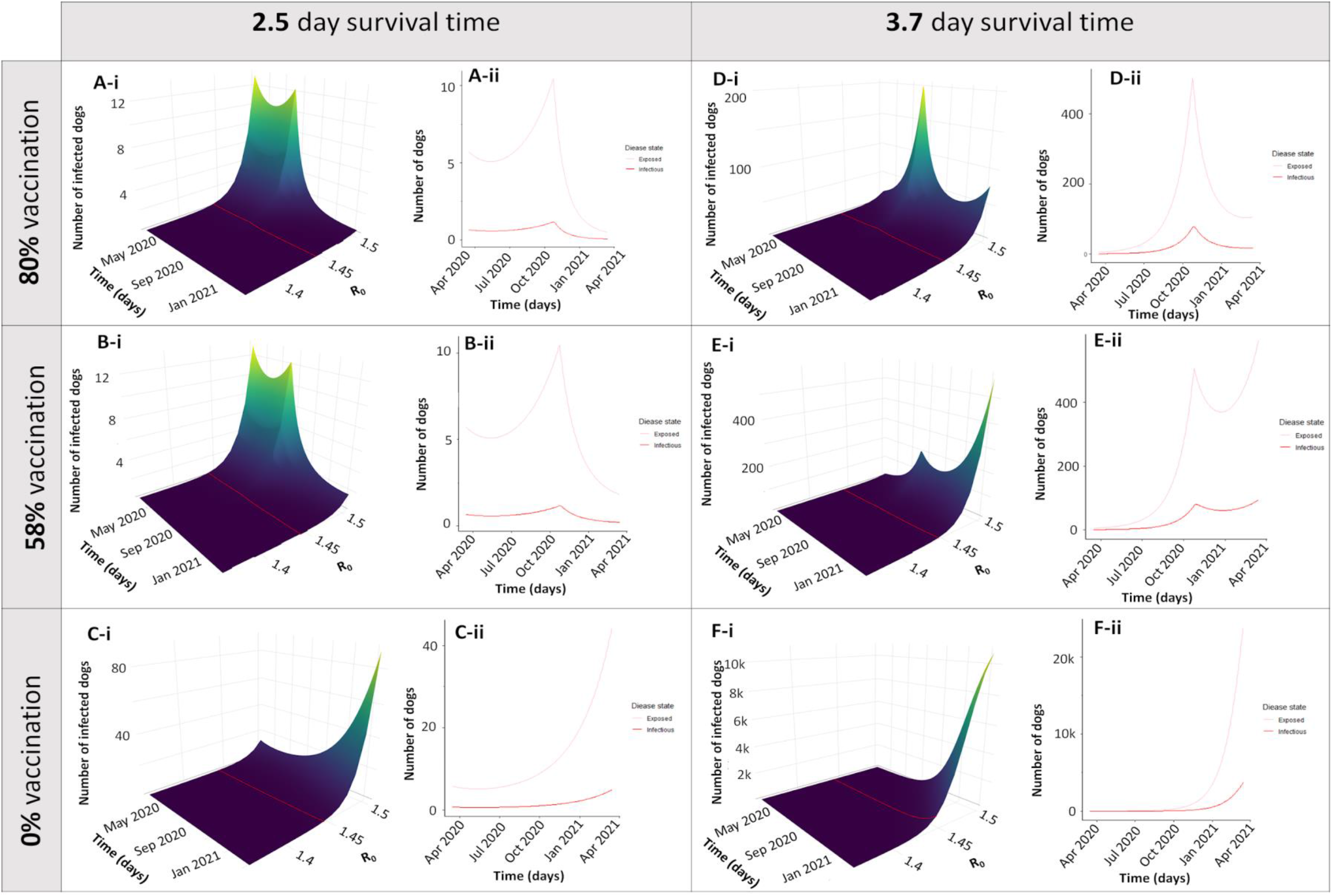
Different simulations of disruption scenarios. Simulations were run for 1 year after the beginning of COVID-19 control measures in Arequipa, Peru (March 16, 2020 - March 16, 2021). Panels A-C depict different vaccination scenarios with normal levels of surveillance and control measures leading to an average survival time (ST) of rabid dogs to be 2.5 days. Panels D-F show the same vaccination scenarios with decreased surveillance leading to an increased survival time of rabid dogs to 3.7 days. The vaccination scenarios depicted correspond to yearly vaccination campaigns reaching the optimal goal of 80% coverage (Panels A, D), a sub-optimal level of 58% coverage (Panels B, E), and a complete cancellation of the vaccination campaign were coverage is 0% (Panels C, F). Both the surface plots with a range of values of R_0_ (i) and a transect where R_0_=1.44 (ii) are displayed)

## Discussion

Our results show that stopping or pausing dog vaccination and rabies surveillance during the COVID-19 pandemic will substantially increase the number of cases of canine rabies with an associated increased risk of human rabies within a few months. The lockdowns have the potential to reduce contact between rabid dogs and humans, but essential activities and some measures to reactivate the economy require people to leave their houses in a city with the highest rate of dog bites in Latin America (*42*). This model is a logical tool to clarify trends that may result from neglect of rabies due to COVID-19 (as opposed to a tool to predict exact numbers of infected dogs on specific dates). We examined COVID-19 caused disruptions of mass dog vaccination campaigns, surveillance activities, and focus control over a wide range of plausible R_0_ values. Though the magnitude of the number of infected dogs changes with R_0_, the trends show that decreasing vaccination coverage and increasing survival time of infectious dogs due to decreased surveillance and focus control will result in increased rabies burden in Arequipa.

This analysis of the city of Arequipa has broad implications regionally for Latin America. Our model of canine rabies in Arequipa demonstrates the effects of COVID-19 on the spread of canine rabies in a city with a medium to large human population (about 1 million), active immigration and emigration, continuous but suboptimal efforts to control rabies, and a fairly large free-roaming dog population; many of these characteristics are shared with other Latin American urban areas. Also, Arequipa represents an area of rabies reintroduction and rabies re-establishment, both undesired and rare events. Modelling a city with these characteristics and continuous rabies virus transmission may provide insight into rabies-endemic Latin American cities during the COVID-19 pandemic and also into the risk of expansion to non-endemic neighboring cities.

COVID-19 has had devastating effects world wide. Case and death counts are the most obvious manifestation of this pandemic. The economic ramifications due to lock down and quarantine have been felt keenly as unemployment has increased and businesses have struggled. However, a more insidious effect of major health crises, including SARS-CoV-2, SARS-CoV-1 (*43*), Ebola (*44*), and Zika (*45*–*47*) is the cost to other public health measures through necessarily diverted funds, resources, and attention. In the past seven months, hospital intakes have decreased for diseases other than COVID-19 and vaccination programs are behind in administering lifesaving preventatives (*48*–*51*). Rabies prevention program disruptions are one of many public health initiatives that have the potential to cause future problems due to COVID-19 induced neglect.

In rabies-affected areas, the COVID-19 pandemic has the potential to interrupt the multi-pronged rabies control program that Peru has developed over recent decades at several points. The first prong of Peru’s rabies control program that may be affected by COVID-19 is the rabies surveillance system. Because Peru’s surveillance system relies heavily on submission of samples from dogs reported to exhibit signs of rabies by the public, the absence of people leaving home to observe these dogs has caused greatly decreased reporting rates. Since COVID-19 control measures began in March 2020, there has been a 10-fold decrease in samples submitted to be tested for rabies. Furthermore, lockdowns have hindered the ability of public health teams to respond and remove rabid dogs. Rabid dogs are often killed by motor vehicles because they behave erratically and run into traffic (*40*). With the government-mandated quarantine and associated decrease in travel, there are fewer cars on the streets and fewer people outside to notice erratically behaving dogs. For these reasons, we postulate that dogs may live longer and transmit rabies to a larger number of dogs before they die, as reflected by a decreased parameter *α*, the death rate due to rabies. From our focus control data, we find that dogs are removed an average of 2.5 days after becoming symptomatic though these data do not reflect traffic deaths. Our model indicates that an increased average survival time of 1.2 days (consistent with dogs dying of rabies as opposed to being killed) could cause a rise in rabies cases. Parallels of decreased surveillance measures can be drawn to the ebola crisis, where protocols to contain ebola interrupted screening and diagnosis of malaria, tuberculosis, and HIV (*52*, *53*). Adequate surveillance is one essential piece of infectious disease control programs that is often neglected for more immediate emergencies.

The second prong of rabies control in Arequipa is yearly vaccination campaigns. Due to COVID-19, the yearly dog vaccination campaign in Arequipa is planned to be reduced, delayed, and possibly discontinued this year, and similar disruptions are expected across Latin America. COVID-19 poses challenges to rabies vaccination campaigns in a few ways. First, geographic areas with high rates of canine rabies in particular need of vaccination points also tend to be areas with high rates of COVID-19 due to population density, which leads to concern for the safety of healthcare personnel and dog owners. Second, public health organizations are focusing their energy and resources on the COVID-19 crisis at hand. Diverting scarce public health resources towards a crisis is often necessary for some amount of time; however, our model suggests that indefinitely postponing vaccination campaigns could have detrimental consequences on the spread of dog rabies and, ultimately, public health. In many areas of the world, diverting public health resources to COVID-19 has resulted in the suspension of childhood vaccination campaigns in many low- and middle-income countries. Eighteen of the 29 countries that have had to pause their vaccination programs as a result of lack of resources or supply chain breakdown are already reporting outbreaks of measles, a highly-infectious disease usually prevented by the MMR vaccine (*54*, *55*). This pattern of upticks in diseases pushed out of focus by public health crises occurred during the ebola epidemic, when national malaria control programs in West Africa dissolved and influenced the increased morbidity and mortality from the disease (*44*). Even if vaccination programs are able to continue on a national scale, vaccination rates have dropped during the COVID pandemic due to the avoidance of medical facilities. In the United States, the number of non-influenza childhood vaccines provided by its Vaccines for Children Program decreased by approximately 3,000,000 doses compared with the same week a year prior and the number of measles-containing vaccines decreased by approximately 400,000 doses (*56*). These steep declines are likely due to adherence to government stay-at-home orders (despite medical recommendations to continue attending well visits) as well as caregivers’ fears of contracting COVID-19 at healthcare facilities (*56*). Our model indicates that though the recommended 80% vaccination coverage goals may be unattainable, an intermediate effort can still have a tremendous effect in curbing the rise of canine rabies

The model presented above has many sources of uncertainty, perhaps the most significant is a likely massive underreporting of canine rabies cases leading to lack of accurate data and bias in parameterized data around which the model is constructed. Inadequate surveillance can exacerbate underreporting— a common problem in rabies-affected areas (*3*, *6*, *10*, *13*, *29*, *32*)—and lead to more cases as the virus spreads undetected. Furthermore, in Peru, one of the COVID-19 control measures is to shut down markets where more sellers are seropositive. These markets usually attract free-roaming dogs and provide them with food (organic trash). Rabies surveillance professionals have noticed that after shutting down the largest markets in Arequipa dogs are wandering farther to find food. The impact of these changes in dog behavior is unknown. The COVID-19 pandemic likely has caused many changes in both human and dog behavior not captured explicitly in our model. These effects are compounded by the limitations of our model; it is a deterministic model and does not capture the changeable nature of outbreaks. Next steps include building a more flexible, stochastic model around the current framework.

Our model demonstrates that the effects of stopping or pausing rabies prevention activities could have serious effects on future cases of canine rabies, and consequently, on the risk of human rabies. Given that COVID-19 will continue to challenge public health departments in the short- and medium-term, it is essential to create a strategy for rabies surveillance and prevention during the COVID-19 pandemic. This strategy should consider new approaches to dog vaccination that can accommodate social distancing and other COVID-19 prevention measures. New dog vaccination approaches, even with suboptimal coverage, could prevent canine rabies cases in the short term. However, an uninterrupted optimal level must be reached for the long-term goal of elimination, as has been shown previously (*57*, *58*). The epidemiological model presented in this paper indicates that decreasing or stopping rabies programming during the pandemic could have downstream effects on rabies in Peru, and likely in the region, and even threatens to undo the remarkable achievements in decreasing rabies cases over the past several decades. This outcome would join a number of other disease spikes that occurred following natural disasters or public health crises. However, it is an outcome that can be avoided with a proactive and careful approach to balancing COVID-19 prevention with rabies surveillance and vaccination programming.

## Data Availability

Parameters were extracted from literature or obtained from analyses of secondary data. Secondary data are available at the Peruvian Ministry of Health upon request.

## Acknowledgement

We gratefully acknowledge the contributions of and the work done by the Gerencia Regional de Salud de Arequipa, the Red de Salud Arequipa Caylloma, the Laboratorio Referencial Regional Arequipa, and the Centro de Salud de Alto Selva Alegre. We acknowledge the work of the members of the Zoonotic Disease Research Laboratory, One Health Unit, and their contribution collecting part of the data used in this study. Dr. Castillo-Neyra was supported by NIH-NIAID grant 1K01AI139284. The funders had no role in study design, data collection and analysis, decision to publish, or preparation of the manuscript.

